# Evidence to inform effective alcohol pricing policies in the European Union

**DOI:** 10.1101/2022.07.28.22277988

**Authors:** Colin Angus

## Abstract

**Aim:** To map current alcohol pricing policies across the European Union and United Kingdom and review the latest evidence on their effectiveness

**Design:** Current policies were mapped using publicly available data. Evidence was systematically reviewed using a three-stage approach: a) a systematic search for published studies in PubMed and Google Scholar, b) a snowball search of grey literature and hand-searching the references of existing reviews and c) consultation with topic experts.

**Setting:** Any appraisal or evaluation on the impact of an alcohol pricing policy in an EU or UK nation that reported alcohol consumption or health outcomes.

**Participants:** The general population

**Measurements:** Any reported measures of alcohol consumption or alcohol-related health outcomes.

**Findings:** The mapping exercise found that there is substantial variation in both the levels and structures of alcohol taxation across Europe. The review found 83 studies, consisting of 34 prospective modelling studies and 49 retrospective evaluations. These came primarily from the UK and Scandinavia. The majority of studies looked at the impact of changes to alcohol taxation, although a substantial minority looked at the impacts of Minimum Unit Pricing for alcohol. Studies consistently fond that increases in taxation, or the introduction of Minimum Unit Pricing, have led to reductions in alcohol consumption and improvements in public health and, in spite of concerns about cross-border sales moderating these benefits, there is little evidence to support these concerns in practice.

**Conclusion:** There is ample evidence to show that alcohol pricing policies can and have worked across Europe and are likely to form a key part of any effective policy approach to reduce alcohol-related harm.

## Background

Alcohol consumption places a substantial burden on the health of European society, causing an estimated 584,000 deaths (6% of all deaths) and the loss of 21 million Disability-Adjusted Life Years every year across the WHO European region (1). In addition to this, excessive alcohol consumption is also responsible for increasing crime and public disorder (2), reducing economic productivity (3) and causes significant harms beyond those suffered by the drinkers themselves (4). Overall, alcohol consumption in Europe has fallen in the past 20 years, however it remains the highest in the world and research suggests that the recent decline is expected to slow in coming years (5).

Stakeholders wishing to address high levels of alcohol consumption and associated harm are not without policy tools at their disposal. There is an overwhelming body of research evidence, running to hundreds of individual studies, which have demonstrated that increasing the price of alcohol is an effective means of reducing alcohol consumption (6–9). As a result, alcohol pricing is listed among the World Health Organization’s ‘best buy’ interventions for reducing alcohol-related harm (10). A previous research project, the AMPHORA study, reviewed the scientific evidence specific to Europe in relation to alcohol pricing policies in 2011 and concluded that “The accumulated knowledge base tells us that restrictions on the physical and economic availability of alcohol have a significant effect on reducing alcohol consumption and related harms” (11).

However, not all alcohol pricing policies are equal. The umbrella term of ‘pricing policies’ covers a broad range of specific policy ideas, which may have different magnitudes of effect in different countries and contexts. Furthermore, the extent to which the impacts of different pricing policies are distributed equally across the population, or whether they effectively target specific groups of drinkers, may vary. The question of whether alcohol pricing policies can effectively target heavier drinkers without having a substantial impact on those who drink within national drinking guidelines has become an important political consideration in many countries. There is also a substantial body of evidence which shows that the burden of alcohol-related harm falls disproportionately on the most deprived parts of society (12), and thus the question of whether pricing policies can reduce the resulting health inequalities has also featured heavily in many political debates around alcohol policy in recent years.

By far the most common pricing policy in place across the European Union is alcohol taxation, with every Member State levelling some form of duty on alcohol (13). As a result, the studies identified and reviewed in the AMPHORA study were almost exclusively ones that assessed the impact of changes in alcohol taxation on a range of outcomes, including alcohol consumption, alcohol-related hospital admissions and mortality. In more recent years, however, there has also been significant scientific and political interest in another pricing policy – Minimum Unit Pricing (MUP). MUP sets a floor price below which a fixed volume of alcohol (e.g. a standard drink or ‘unit’) cannot be sold. Variations of MUP have been in place in several Canadian provinces for many years, but the last decade has seen a flurry of research and policy interest in MUP within EU Member States^1^. This has been largely driven by Scotland, which passed legislation to introduce MUP in 2012 and finally brought the policy into force in 2018 following a lengthy legal challenge from the alcohol industry (14). The past decade has also seen developments in other forms of pricing policies, with Scotland introducing a ban on multi-buy discounts (promotions where consumers pay a cheaper per-unit price for buying larger volumes of a product) for alcohol in 2011. Finally, there has also been an increasing focus on not only the *levels* at which alcohol taxes are set, but also in the *ways* in which they are levied (15,16).

In light of these important developments in alcohol pricing policy, it is important to understand how the scientific evidence has moved on in recent years in order to inform the development of more evidence-based alcohol pricing policies across the EU region. To this end, we set out to achieve the following aims:

1. To map current alcohol pricing policy across the European Union
2. To identify and review the latest research evidence on alcohol pricing policies in the European Union
3. To consider relevant international research evidence on alcohol pricing policies
4. To synthesise these findings to make recommendations about the potential impacts of different alcohol pricing policies in EU Member States

These aims will be achieved through a combination of updates of previous reviews and analyses, a systematic review of published scientific studies and through consultation with leading topic experts.

## Mapping current pricing policies across the EU

### Methodology

We updated previous analyses of EU alcohol duties (13,16) using the latest available EU-wide data (17) and combined these with wider data on alcohol production (18).

### Findings

There are three main approaches to levelling alcohol taxation (15):

1. Duty based on the volume of the product (unitary taxation)
2. Duty based on the volume of alcohol contained in the product (specific taxation)
3. Duty based on the value of the product (ad valorem taxation)

Within the EU, regulations on the harmonisation of alcohol duties require that beer is taxed on a specific basis (except under specific circumstances detailed in (13)), wine is taxed on a unitary basis, with differential rates permitted for products below 8.5% ABV, and spirits are taxed on a specific basis. Member States are permitted to levy additional ad valorem taxes, but in practice these are normally general sales taxes such as VAT, rather than alcohol-specific taxes (19). There are also minimum duty rates for both beer and spirits, but no minimum rate for wine (20).

In spite of these restrictions, there is substantial variation in both the levels and structures used to tax alcohol in EU Member States, as illustrated in Figure 1. Beer duties are generally lower, per gram of ethanol, than duties on either wine or spirits and uniquely, the UK has a higher rate of duty for high-strength beer (above 7.5% ABV).

**Figure 1.**
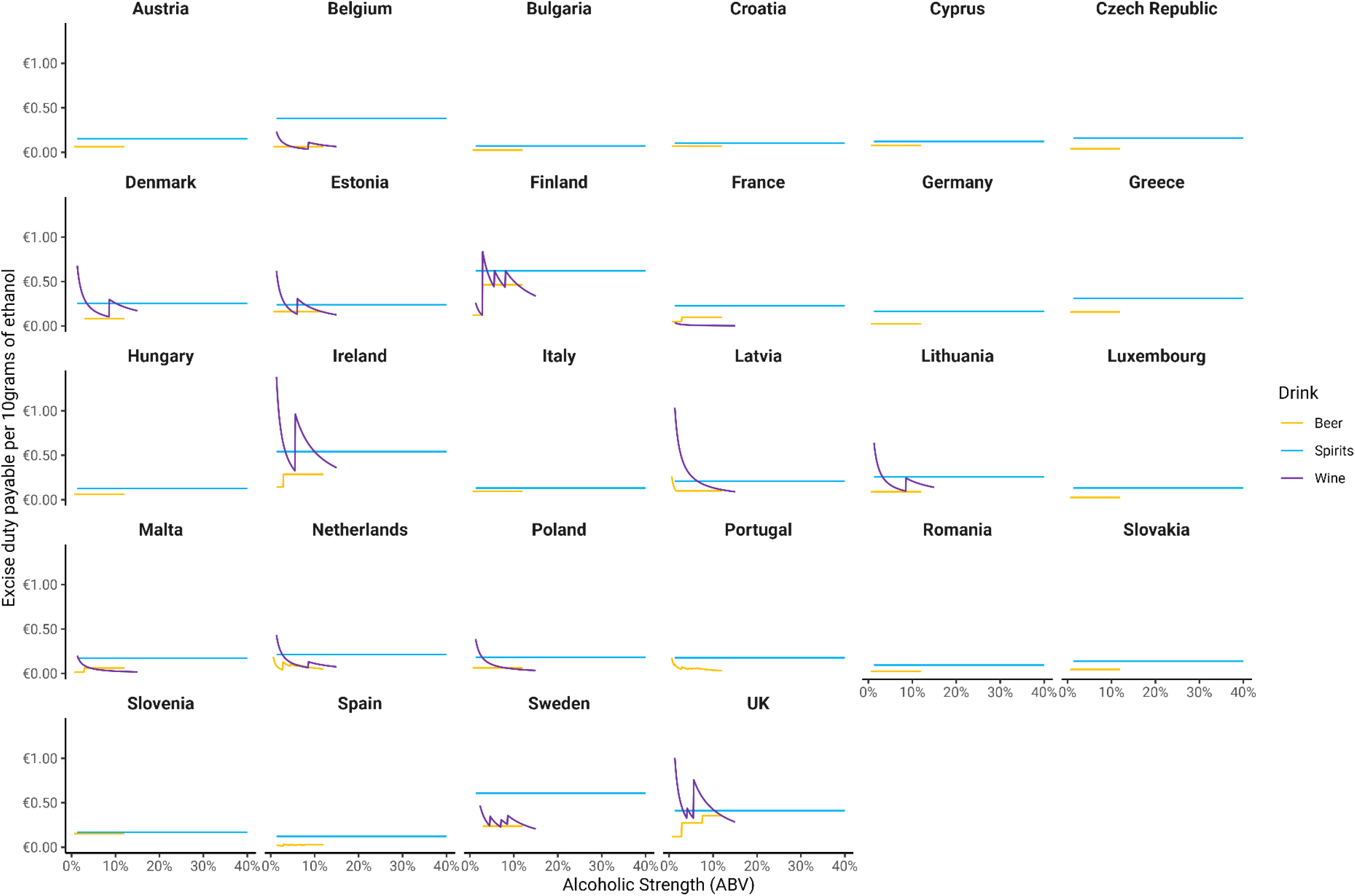
Alcohol duty rates per 10 grams of ethanol by alcoholic strength and beverage type across the EU. Zero duty rates not shown.

The requirement to tax wine on a unitary basis means that the effective duty rate per gram of ethanol falls as strength increases, effectively incentivising consumers to purchase higher strength products. Some Member States have partially addressed this through the use of differential duty bands (particularly Finland and Sweden), although the fact that duty rates must remain fixed between 8.5% and 15% ABV, the range in which most wine products lie, means this is likely to have limited impact.

Taxes on spirits are almost universally higher than on other alcoholic products, perhaps motivated by the fact that spirits are more closely associated with intoxication, as they allow a greater volume of alcohol to be consumed in a shorter time.

For all products there are large differences in alcohol duty rates between countries, with the duty payable on 500ml of 5% ABV beer varying between 5c in Bulgaria, Spain, Luxembourg, and Romania and €0.91 in Finland. The duty levied on a 700ml bottle of 40% ABV spirits ranges from €1.57 in Bulgaria to €13.66 in Finland, while 15 Member States would levy no duty on a 750ml bottle of 12.5% ABV wine compared to €3.19 of duty in Ireland.

If we look at those countries which do not levy duty on wine, there is a striking pattern, shown in Figure 2, when we compare duty rates with data on per capita wine production (18). All major wine producing nations charge no duty on wine, while Member States who produce little or no wine of their own generally have non-zero rates of duty on wine.

**Figure 2.**
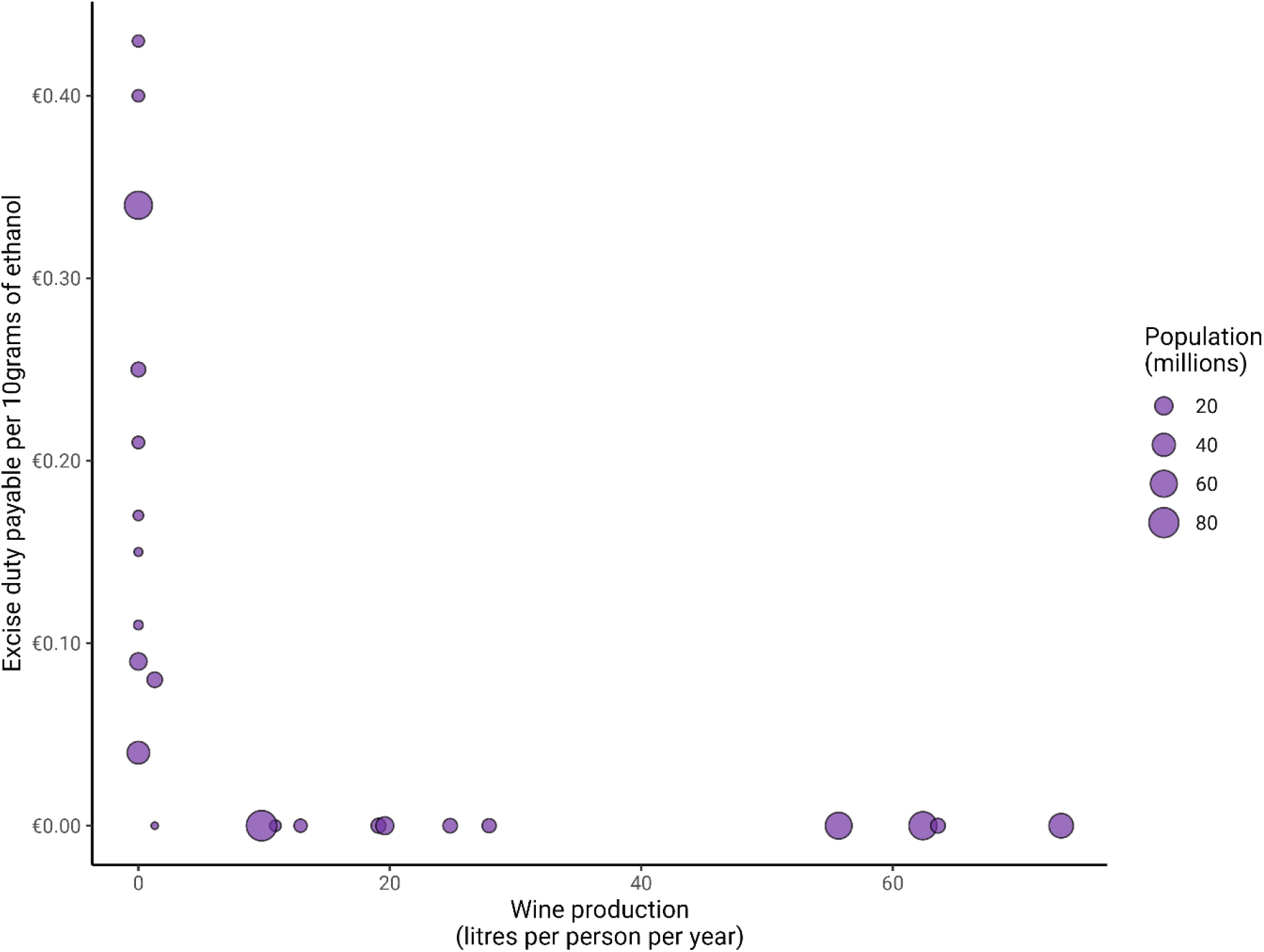
A comparison of per capita wine production and duty rates levied on 12.5% ABV wine for EU Member States

**Figure 3.**
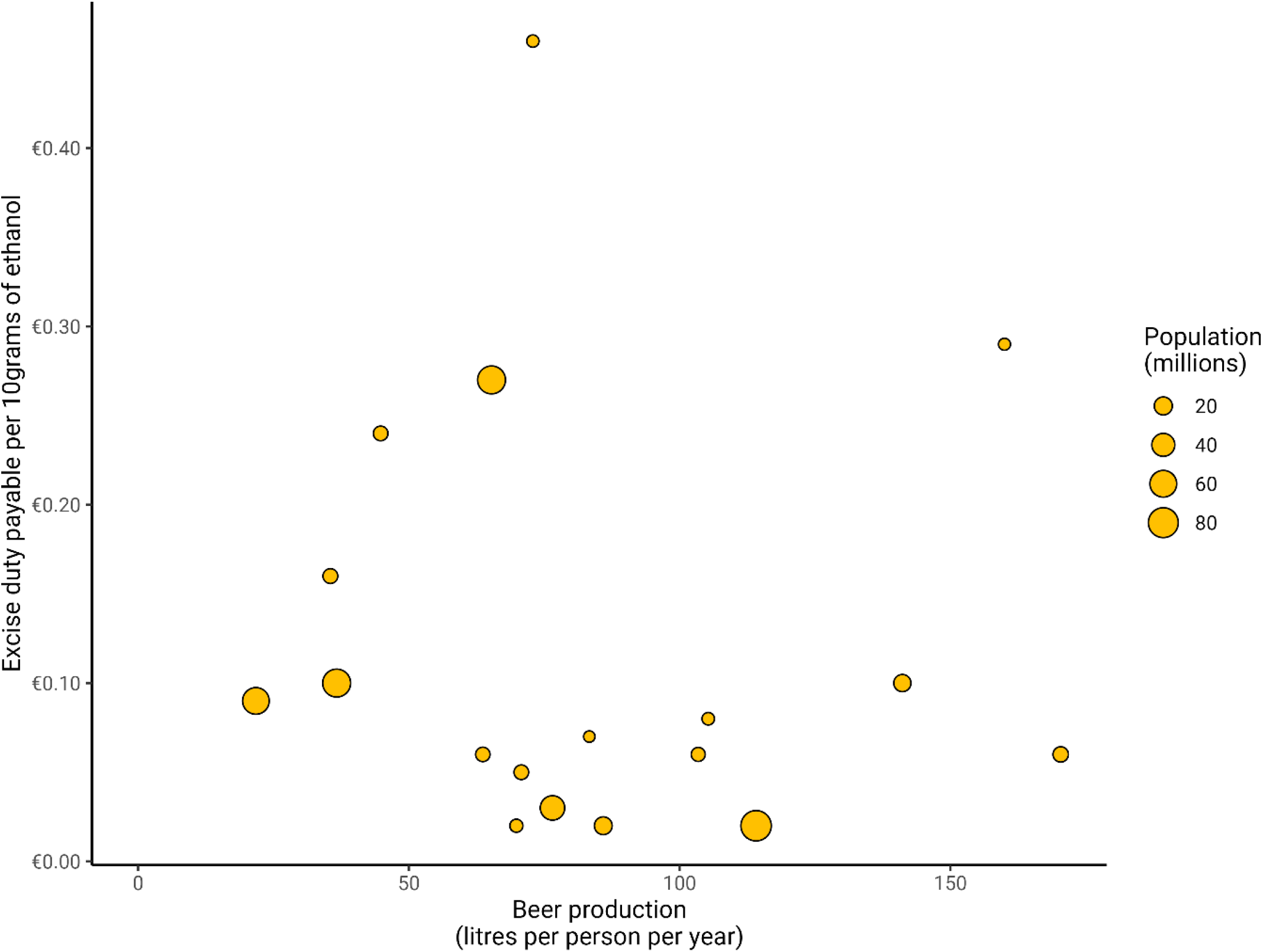
A comparison of per capita beer production and duty rates levied on 5% ABV beer for EU Member States

While this pattern may suggest some degree of protectionism, the picture for beer production is very different, with no significant association between per capita beer production and rates of duty levied on beer. This contrast would appear to call into question any argument that low rates of duty are necessary to support a country’s own alcohol producers.

Beyond alcohol duties, Scotland introduced a Minimum Unit Price of 50p per UK unit (equivalent to €0.73 per 10g of alcohol) in May 2018. Wales followed suit in March 2020 and Ireland have recently confirmed that they will bring in a MUP of €1 per 10g of alcohol at the start of 2022. Scotland introduced a ban on multi-buy discounts in 2011, a policy that Finland and Sweden also have in place. Finally, England and Wales have had a ban on selling alcoholic products for below the cost of the duty levied on the product plus VAT since 2014. Several other EU countries have wider regulations preventing the sales of many common products, not just alcohol, for below the retailers’ own purchase costs.

## Reviewing published studies on alcohol pricing

### Methodology

We sought to identify studies which evaluated or appraised the impact of an alcohol pricing policy on either alcohol consumption or health, in an EU Member State, which had been published since the year 2000. Studies which assess associations between alcohol prices and outcomes without specifically looking at the impact of a pricing policy (e.g. econometric studies estimating price elasticities) were excluded.

We took a three-pronged approach to identify relevant studies:

1. A systematic review of published academic studies
2. A search through grey literature and the references of existing reviews
3. Consultation with topic experts

For the systematic review, we searched the PubMed and Google Scholar databases for any study (including grey literature) which met the inclusion criteria outlined above^2^. We then hand searched the references of existing published reviews of alcohol pricing policies (21–26) and the references and studies citing key papers identified in the systematic review. We then consulted with key topic experts to ensure we had not missed any important studies.

For all identified studies, we extracted bibliographic details, the country (or countries) under study, the pricing policy in question, the study type, the intervention period, the outcomes being examined (including any non-health outcomes reported alongside consumption or health) and the study findings.

### Findings

The initial systematic review identified 11,623 studies, which were reduced to 108 after screening title and abstract and leading to a final total of 41 eligible studies after full text screening. These were supplemented with a further 42 studies identified through the additional searching and expert consultation phases to give a total of 83 studies. These are summarised narratively below

#### Study types

The identified studies can be grouped into two broad categories:

- **Prospective modelling studies** which use mathematical models to estimate the potential impact of policies which have not yet been introduced
- **Retrospective evaluations** which use a variety of methods such as Interrupted Time Series analysis to evaluate the impact of a single intervention which has already occurred on a specific outcome

We identified 34 prospective modelling studies and 49 retrospective evaluations. The majority of included studies of both types were published in the last decade, as illustrated in Figure 4.

**Figure 4.**
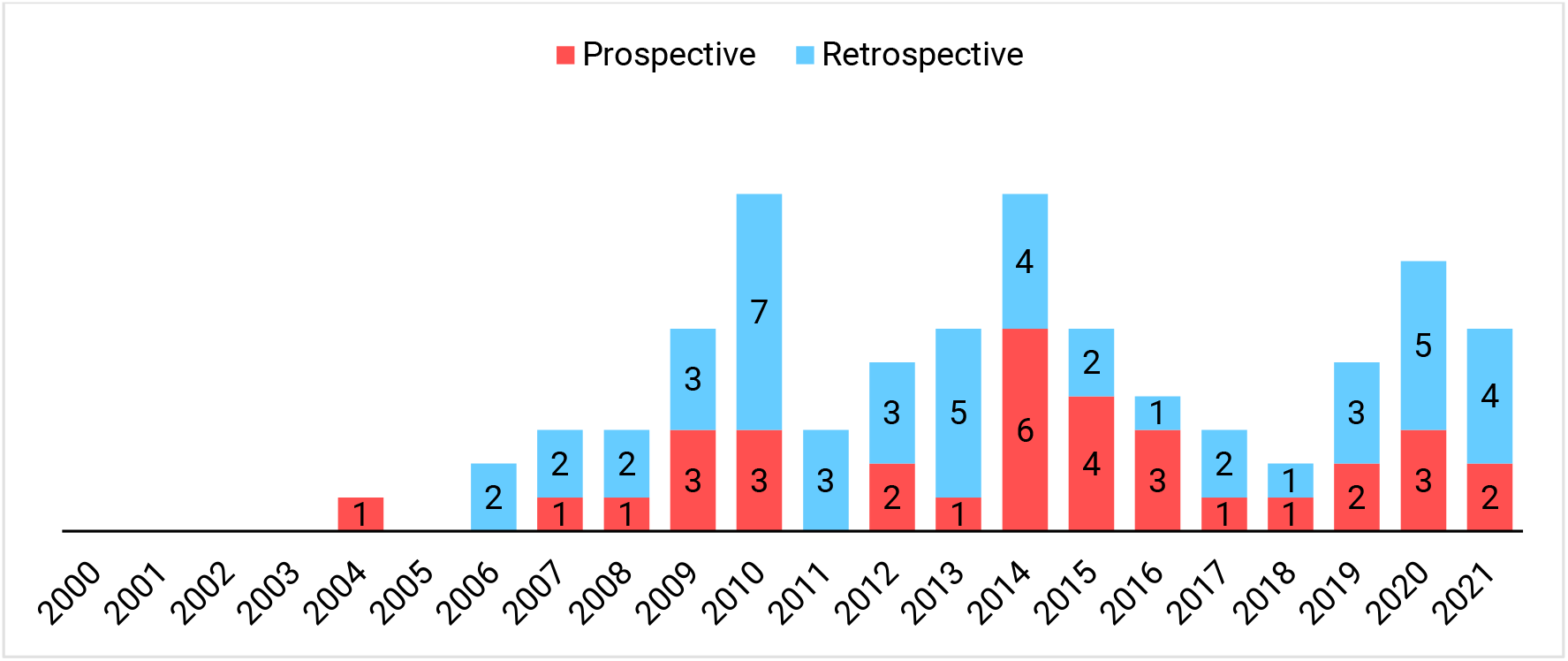
Publication date of included studies

#### Study location

The country setting for all included studies is shown in Figure 5. There is a heavy bias towards Scandinavian countries and the UK in the settings of the identified studies, with 24 of the prospective studies (71%) and 42 of the retrospective studies (86%) assessing the impact of interventions from these countries. It may be tempting to conclude that this is because these countries have seen more alcohol pricing policies implemented or considered during this period. While that may (or may not) be true, the AMPHORA study identified a large number of unevaluated pricing policy interventions from other parts of Europe (27), suggesting that policies from Southern and Eastern Europe are less likely to be studied than those in the UK or Scandinavia.

**Figure 5.**
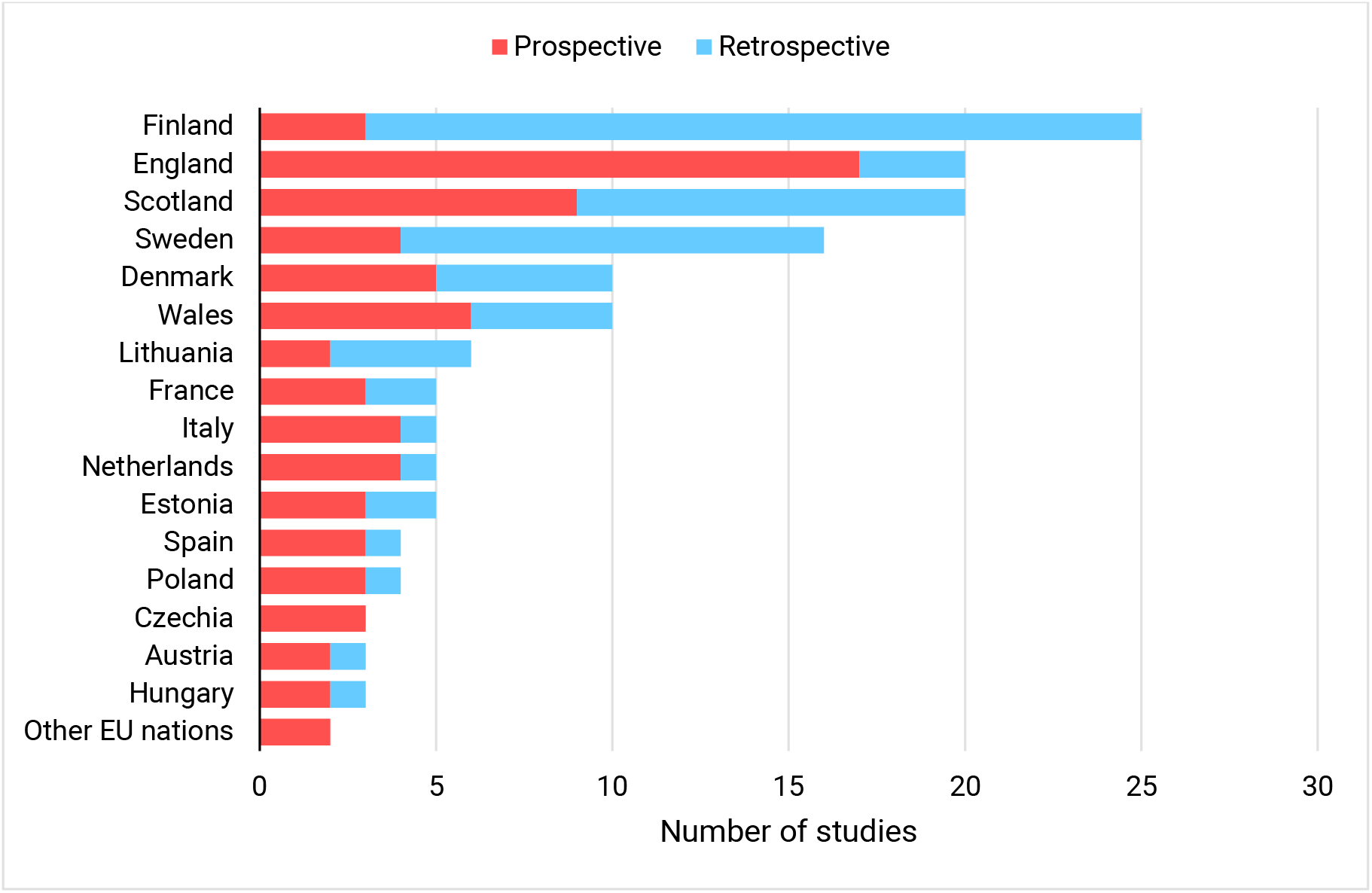
Identified studies by country and study type

#### Intervention type

The nature of the pricing policies examined in the included studies is presented in Figure 6. This shows that studies of taxation policies are dominant, but that there are a significant minority of studies around Minimum Unit Pricing. There is also a clear tendency for retrospective studies to look at tax changes, while prospective studies have looked at MUP to a greater extent.

**Figure 6.**
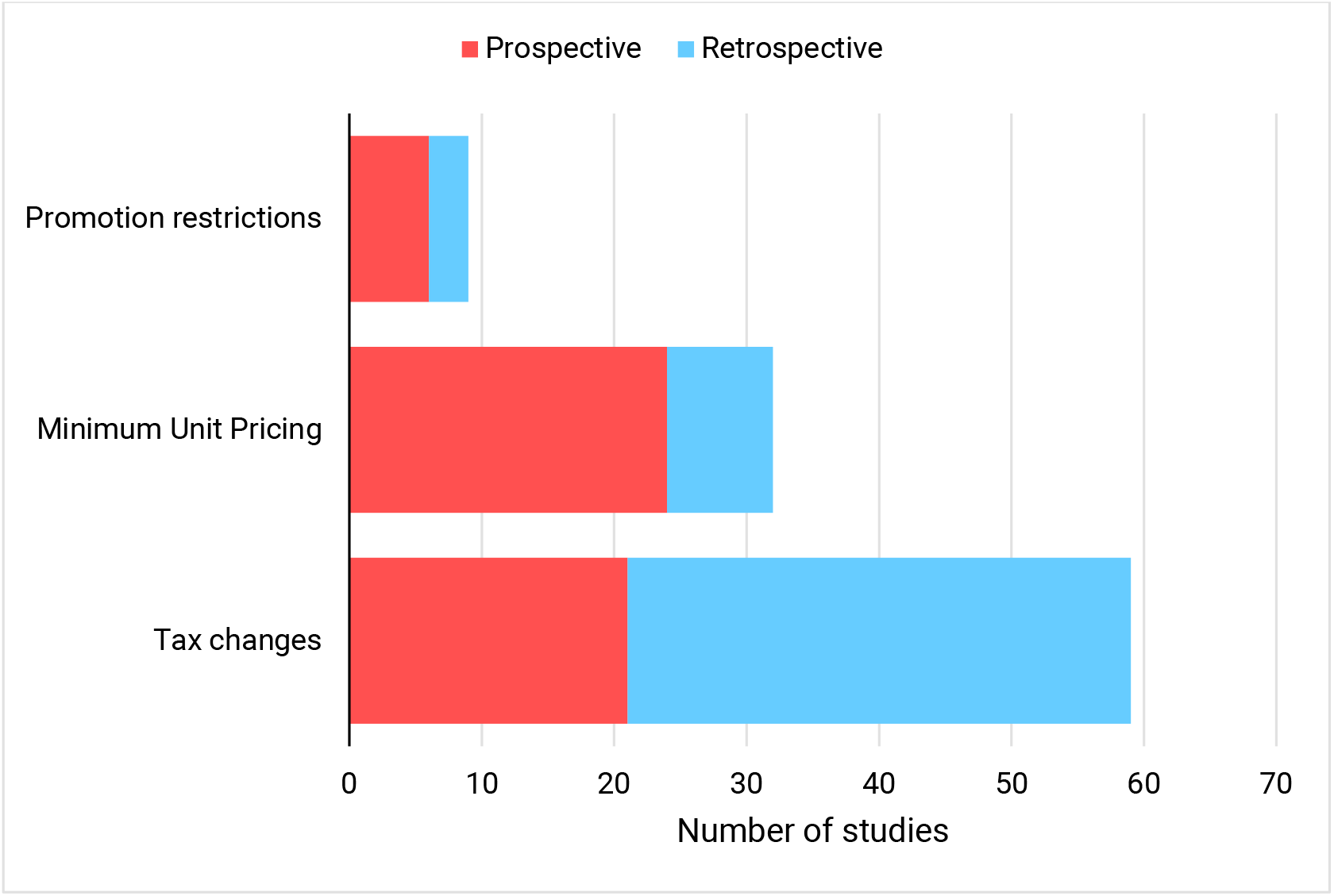
Types of alcohol pricing policy examined in included studies

#### Prospective modelling studies

The 34 prospective modelling studies identified in the review cover a wide range of methodological approaches. Several studies take a broad approach using aggregate data to estimate the impact of multiple policies across multiple countries or regions (28–32). The remaining studies take a more detailed approach, using low-level individual data on how alcohol consumption and harm vary within the population and modelling the impact of policies on different subgroups in the population, although not all of the studies report the subgroup-level impacts (33–61). The studies include a wide range of outcomes, from alcohol consumption and spending to alcohol-related hospital admissions, deaths, crimes and workplace absence. It is striking that across all of these outcomes in all of the included modelling studies, the findings are clearly positive, showing that alcohol pricing policies are effective, cost-effective and health-improving.

Among the more aggregate studies, Chisholm et al found that increasing alcohol taxes would improve health across all WHO Europe subregions (30) and Summan et al estimated that a 20% increase in alcohol duties would reduce the number of Years of Life Lost to premature mortality and increase tax revenue for countries at all income levels around the globe (28). Rovira et al found that increasing alcohol taxes would significantly reduce alcohol-attributable cancer cases in Germany, Italy and Sweden (29), while Lai et al estimated that an increase in alcohol taxes would improve population health and be highly cost-effective in Estonia (31). Two other studies looked across multiple countries, concluding that increasing alcohol prices would reduce chronic disease across 11 EU Member States (32) and that both increasing taxation and introducing an MUP would reduce alcohol consumption, improve health and reduce healthcare costs in Czechia and Germany (44).

Two separate studies led by Holm appraised the cost-effectiveness of multiple alcohol policy interventions, including increased taxation, in Denmark and concluded that increasing alcohol taxes would improve health and save costs (33,34). A Dutch study compared the modelled impacts of a small increase in alcohol duties compared to increasing them to the same levels as Sweden (a substantial increase), finding that both policies would improve health and be highly cost-effective, although not cost-saving (35). Two separate studies set in Germany appraised the potential impact of increasing alcohol prices on underage drinking (43) and alcohol-related cancer (42), finding that both would be reduced.

22 of the remaining 23 studies use various iterations of the same modelling approach – the Sheffield Alcohol Policy Model, to estimate the impact of a range of policies including Minimum Unit Pricing, tax increases, changes to the structure of the tax system and restrictions on discounting and promotions in England (36–41,45,49,52–56), Scotland (45–48,51), Wales (50,57,58), Northern Ireland (60) and Ireland (59). Of particular note among these studies are the consistent findings across all of these countries that MUP would effectively target the heaviest drinkers while having little impact on the drinking of those consuming within official low risk drinking guidelines and that it would effectively target heavier drinkers on lower incomes, leading to a reduction in socioeconomic inequalities in health.

Several of these studies directly compare the impact of MUP and increases in alcohol taxation (37,46,57), estimating that while both policies would be effective at reducing alcohol consumption and harm, MUP is more effectively targeted at heavier drinkers and more effective at reducing inequalities than tax increases. These differences arise because heavier drinkers tend to consume cheaper alcohol. Although increasing taxation on alcohol increases the price of these products, it also increases the prices of other products to a similar extent. On the other hand, MUP increases *only* the prices of the cheapest products. As a consequence of this, another consistent finding across these studies is that higher levels of MUP are estimated to be more effective, but less targeted at the heaviest drinkers, meaning that they have a greater impact on the drinking of moderate consumers. One of these studies looked further at alternative approaches to taxation, finding that a purely specific tax system would be almost as effective as MUP at reducing health inequalities (37).

One study compares the impact of MUP to the impact of introducing a ban on selling alcohol at below the cost of the tax levied on the product, a policy which was subsequently introduced in England and Wales (39). This study concluded that while banning sales below cost was unlikely to be harmful, it was estimated to be 40-50 times less effective than introducing MUP instead. Another study looked at gendered differences in the impact of pricing policies, concluding that both duty increases and MUP are likely to have greater impacts on the drinking and subsequently on the health of men than on women (40).

The final study, conducted by Griffith and Smith, compared the impact of MUP to a reform of the taxation system, which included a switch to a purely specific tax system (i.e. all products being taxed on the basis of their alcohol content) (61). This research estimated that the alternative tax system would be almost as effectively targeted at heavy drinkers as MUP, but would have the benefit that the policy would increase revenue for the government through increased duty receipts, whereas the majority of the increased sales value under MUP is estimated to go to retailers and producers.

#### Retrospective evaluation studies

As the 49 included evaluation studies include multiple studies evaluating the impact of the same intervention, we will group these studies by country and intervention. Table 1 gives an overview of the characteristics of the studies identified in the review.

**Table 1.**
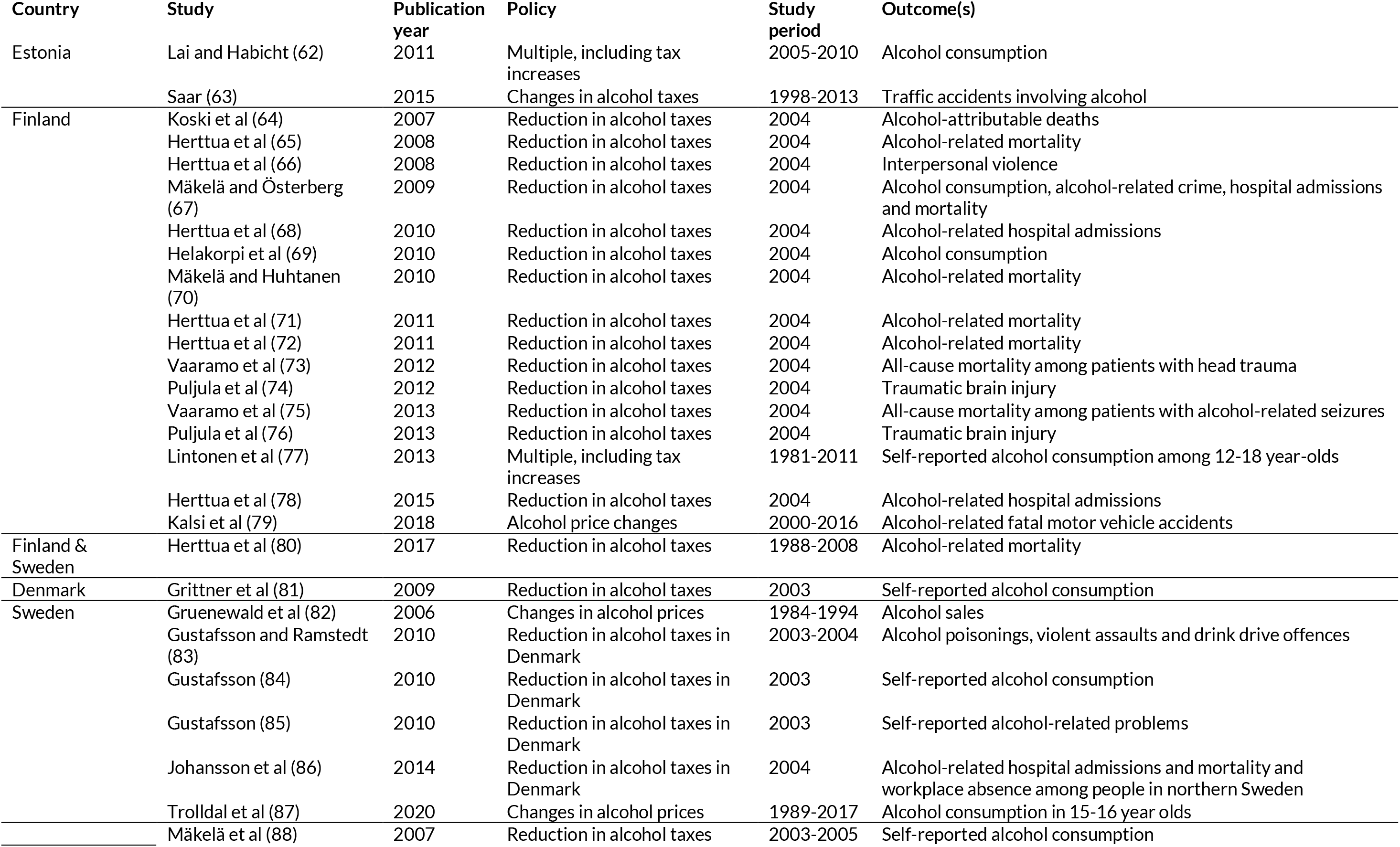
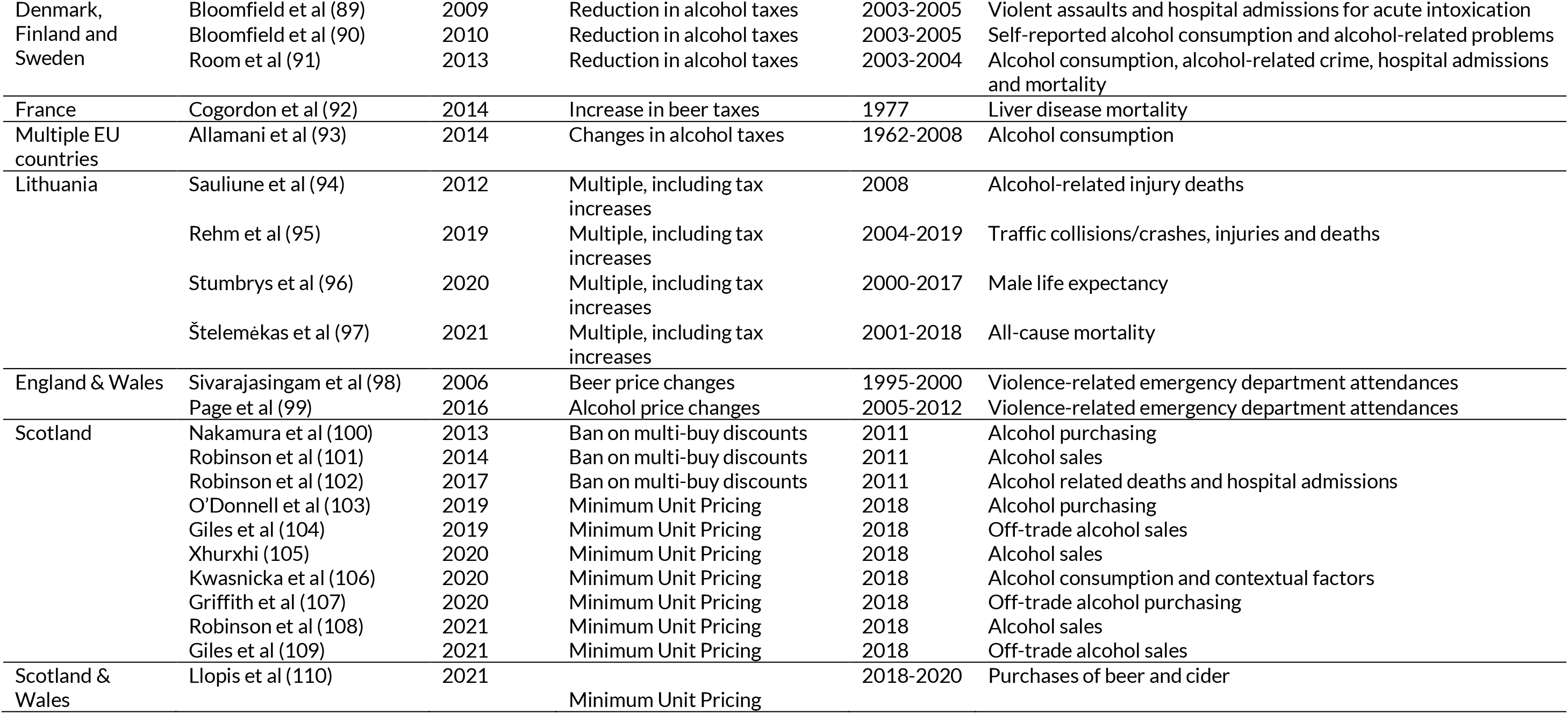
Summary of included retrospective evaluation studies

##### Estonia

Alcohol taxes in Estonia were increased between 2005-2010, alongside a number of other alcohol control policies. One short-term study found an association between the introduction of these policies and a fall in alcohol consumption (62), although this cannot be attributed to any one policy. A second study looked over a longer time frame and found a significant inverse association between alcohol tax rates and the number of traffic accidents recorded as involving alcohol (63).

##### Finland

In response to neighbouring Estonia joining the EU, Finland cut alcohol taxes by around a third in 2004, fearing significant cross-border shopping. A series of studies have assessed the impact of this tax cut on a wide range of outcomes, finding that it was associated with a significant increase in alcohol-related deaths in the general population (64,70) and that this increase was greater among lower socioeconomic groups (65), those aged 40-70 (71) and those living on their own (72). Other studies found a similar increase in mortality after the tax cuts among patients with head trauma (73) and suffering alcohol-related seizures (75). Other studies found that alcohol consumption, crime and hospital admissions increased (67–69), although there was no evidence of a significant change in rates of interpersonal violence (66) or traumatic brain injury (74,76). Two further studies looked across a longer time frame, with one based on 2000-2016 data finding a significant association between alcohol prices in Finland and alcohol-related fatal motor vehicle accidents (79) and another, which also included data from Sweden, finding weak evidence of a relationship between the affordability of alcohol and alcohol-related mortality (80).

##### Denmark

At a similar time to the Finnish tax cut, Denmark also cut spirits taxes. Evidence from surveys suggests that there was no significant change in the amount of alcohol that people reported drinking following this cut (81).

##### Sweden

An analysis of time series data on alcohol prices and sales using Swedish data found that price increases were associated with significant falls in sales, but that these falls were moderated by drinkers switching to buying cheaper products (82). A similar analysis looking at the relationship between alcohol prices and alcohol consumption in 15-16 year-olds found no significant association (87). Several studies have used data from Sweden to assess whether there was any knock-on impact from the tax cuts discussed above in Denmark and Finland. These studies found a significant rise in alcohol poisonings (83) and in sickness absence from work (86), but no evidence of a change in other health harms (83) or in self-reported alcohol consumption (84) or alcohol-related problems (85).

##### Denmark, Finland and Sweden

Three further studies looked across both the Danish and Finish tax cuts, alongside Swedish data, to evaluate the impacts of these policies across the whole region. These studies did not find any significant evidence of a change in self-reported alcohol consumption or levels of alcohol-related problems (88,90) and little evidence of an increase in harms, although hospital admissions for acute intoxication among the under-16s did rise significantly (89). A further overview study found modest evidence that alcohol consumption and harms increased overall when taxes were cut, but that these effects were not seen across all countries, or in all population groups (91).

##### France/Multiple EU countries

A pair of studies, conducted as part of the AMPHORA project, looked at associations over the period since the 1960s between a wide range of contextual factors, including alcohol policy changes, on liver disease mortality in France (92) and alcohol consumption across a number of EU Member States (93), but did not find any clear evidence on the impacts of pricing policies.

##### Lithuania

In response to high levels of alcohol consumption and related harm, Lithuania introduced a wide range of alcohol control policies in 2007, including an increase in alcohol taxes. This was followed in 2017 by a further series of measures, including more tax rises. One study from 2012 evaluated the impact of the 2007 policies on alcohol-related injury deaths, finding a significant reduction, although this cannot be specifically attributed to the tax increase (94). Three more recent studies also include the more recent policies in their analysis and find that traffic collisions, injuries and deaths (95), alcohol-related mortality in men (96) and all-cause mortality (97) fell when stricter policies were introduced, although again, these effects cannot be directly attributed to the tax increases.

##### England & Wales

Two studies set in England & Wales have looked at associations over time between the price of beer (98) and all alcohol (99) with violence-related emergency department attendances. Both found that higher prices were associated with lower attendance rates, with some suggestion that this relationship was stronger for prices of alcohol in the on-trade (i.e. in pubs, bars and restaurants) than the off-trade (i.e. in shops).

##### Scotland

As part of the 2010 Alcohol Act, from October 2011 the practice of offering multi-buy discounts in retail shops was banned in Scotland. Two studies have evaluated the impact of this policy on alcohol sales, with one finding no evidence of effect (100) and the other finding that sales of wine fell significantly after it was introduced (101). A follow-up study did not find any significant evidence that alcohol-related hospital admissions or deaths changed when the policy was implemented (102).

Subsequently, Scotland introduced a comprehensive MUP policy covering all alcohol sold in all locations in May 2018. This policy has a ‘sunset clause’ whereby it will lapse after 6 years unless the Scottish parliament vote to retain it in law. As a result of this clause, there is a comprehensive evaluation being undertaken of the response to the policy and its impacts, including possible unintended negative consequences (111). There are also a number of independently-funded research studies looking to evaluate other aspects of the impact of MUP. Many aspects of these evaluations are yet to be completed, and will be published in the coming months and years, however to date, 8 studies have been published which meet the eligibility criteria of this review. 6 of these studies have evaluated the impact of MUP on alcohol sales and concluded that overall sales have fallen since MUP was introduced, in contrast with neighbouring England, where alcohol sales have risen over the same period (103–105,107–109). Two of these studies have looked at how consumption has changed differently across the population, finding that the largest reductions in purchasing have come from the households who consumed the most alcohol prior to MUP being introduced, and households on lower incomes (103,107). One additional study used an innovative approach to assess the individual-level impact of MUP on drinking behaviour, finding some tentative evidence for reductions in alcohol consumption among some participants, although the reasons for these reductions appeared to vary between individuals (106).

##### Scotland & Wales

The final study identified in the review looked simultaneously at alcohol sales following the introduction of MUP in both Scotland and Wales, with a particular focus on purchases of low and no-alcohol beer and cider. The study found that overall sales fell in both countries when MUP was introduced and there was some evidence to suggest that consumers had shifted towards lower ABV products at the same time (110).

##### Other relevant studies

In the course of conducting this review, we identified a number of studies which did not meet the eligibility criteria, but which were nonetheless highly relevant to the question of alcohol pricing policies. We will briefly review these papers here.

Connolly et al addressed the question of whether increasing alcohol taxation would negatively impact the economy through a loss of jobs in the alcohol industry using a prospective modelling approach (112). The study estimated that while there may be a small number of job losses within the alcohol industry these would be more than offset by an increase in employment in other sectors.

Ally et al and Wilson et al looked at the extent to which retailers pass on increases in alcohol taxes to their customers in the off-trade (113) and the on-trade (114). Both studies found that retailers do pass price increases through to consumers, but that they do not do so equally across the price spectrum. The cheapest products are generally increased by less than would be expected, offset by larger than expected increases in price for more expensive products. This means that increasing taxes on alcohol is a less effective means of increasing the price of the cheapest products than it would otherwise be.

Lachenmeier et al reviewed the evidence on unrecorded alcohol and concluded that the best approaches to dealing with issues around unrecorded consumption will be specific to the local context depending on the source of the unrecorded alcohol (115).

Two studies from Griffith et al looked at the design of alcohol tax systems and found that a system which uses specific taxation rather than unitary taxes is more efficient and better targeted at heavier drinkers (116,117).

Finally, there have been a number of additional studies published as part of the evaluation of MUP in Scotland. These include a study on compliance which found high levels of compliance among retailers, with no evidence of illegal activity and little suggestion of any cross-border trade (118). Another study looked at the impact of MUP on small retailers, finding some evidence of market adaptations in response to the policy (e.g. some high-strength products reducing their ABV) and also some evidence that products which had been sold at above-MUP levels prior to May 2018 had slightly increased their prices when the policy came in (119). A study from Frontier Economics looking at the wider economic impacts of MUP on the alcohol industry found limited evidence of any negative impact on revenues of retailers or producers, no reports of job losses or reductions in industry investment and limited evidence of local cross-border shopping (120). Finally, a pair of qualitative studies have looked at the impacts of MUP on children and young people, both in terms of their own drinking and the impact on them of the drinking of others (121,122). These found little evidence of any impact (positive or negative) either directly or indirectly on children and young people.

##### International research evidence

The findings of the review align closely with existing studies published in other countries. In particular, the emerging evidence from Scotland on the effects of MUP align with existing studies from Canada which show that increasing the levels of existing minimum prices has led to reduced alcohol consumption (123), hospital admissions (124), mortality (125) and crime (126) and that the largest impacts in terms of reduced harm have been seen in the most deprived areas (127). Initial evidence from Australia’s Northern Territory, which introduced MUP in October 2018 are also looking similarly positive (128,129). There is also an international body of prospective modelling research, which similarly aligns with the studies identified in the review, for example in concluding that MUP policies are likely to reduce health inequalities (130).

## Summary and conclusions

There is substantial variation in levels and structures of alcohol taxation across EU Member States. In spite of overwhelming evidence that increasing alcohol duty rates is an effective approach to reducing harm, alcohol duties remain comparatively very low in many Member States. Evidence from across Europe makes it clear that when Member States have increased alcohol taxes, they have seen benefits, and in spite of concerns about cross-border impacts moderating the effect of increases in taxation, there is little in the published evidence to support those worries.

Current alcohol taxation in the EU is not as effective as it could be, with the requirement to tax wine only on a unitary, rather than specific basis running contrary to public health goals and making it harder for Member States to use alcohol taxation as effectively as possible to reduce levels of alcohol-related harm.

Taxation is not the only alcohol pricing policy. There is limited evidence to suggest that restricting promotions or discounts on alcohol is effective, although it is unlikely to be harmful. There is considerably stronger evidence that Minimum Unit Pricing is an effective, well-targeted policy approach. By changing the price of only the cheapest alcohol MUP can achieve similar overall reductions in consumption and harm to large tax increases while having relatively limited impacts on moderate drinkers. The evidence also suggests that MUP is likely to be more effective at reducing health inequalities. However, MUP is not a silver bullet and the evidence is still emerging, particularly around its real-world effectiveness at reducing alcohol-related harm. Some may also be concerned about the fact that a substantial proportion of the revenue from MUP goes to retailers and producers, rather than government. It may be possible to address this with additional measures introduced alongside MUP, such as a windfall tax on profits, or through a combination of MUP and alcohol tax increases alongside each other.

Ultimately there is no one alcohol pricing policy to rule them all. The ‘best’ pricing policy for any individual situation will depend on the specific local context and also the aims of the policy maker. However, there is ample evidence to show that pricing policies can and have worked across EU Member States and they are likely to form a key part of any effective policy approach to reduce alcohol-related harm.

## Data Availability

All data used in this study is publicly available

## ACKNOWLEDGMENTS AND DISCLAIMER

Part or all of this research was produced under the DEEP SEAS service contract (Contract No. 20177113 - Developing and Extending Evidence and Practice from the Standard European Alcohol Survey - www.deep-seas.eu) with the Health and Digital Executive Agency (HaDEA) acting under the mandate from the European Commission (DG SANTE). The information and views set out in this report are those of the author(s), and the accuracy and veracity of these are the author’s responsibility. Accordingly, the information and views presented during sessions cannot be considered to reflect the views of the Commission and/or HaDEA or any other body of the European Union. The European Commission and the Agency do not accept any responsibility for use that may be made of the information contained therein.

## Footnotes

1 The United Kingdom was still part of the EU when this report was commissioned and therefore is included in this report, even though it is no longer a Member State.

2 The search strategy was based on: (“Alcohol” OR “Ethanol” OR “Wine” OR “Beer” OR “Spirit*”) AND (“Tax” OR “Taxation” OR “Taxes” OR “Price*” OR “Pricing” OR “Economic” OR “Policy” OR “Discount” OR “Promotion*”) AND (“Evaluat*” OR “Apprais*” OR “Model*” OR “Cost*” OR “Cost-effectiveness” OR “Cost-utility” OR “Cost-benefit” OR “Budget*” OR “Value for money” OR “Return on investment”)

